# A Semi-Automated Method for 3D Reconstruction from Routine Ultrasound Videos: Visualization of Transient Gallbladder Lesions

**DOI:** 10.1101/2025.11.11.25340033

**Authors:** Taiki Kojima

## Abstract

**Purpose:** Ultrasound imaging of the gastrointestinal tract faces two major limitations: (1) lesions are often visible only momentarily, and (2) the lack of three-dimensional context makes it difficult to understand the orientation and anatomical location of the observed cross-sections. We aimed to develop a simple, semi-automated three-dimensional (3D) reconstruction method from routine ultrasound videos to enhance the visualization of transient lesions, particularly in the gallbladder.

**Methods:** Ultrasound videos of the author’s own gallbladder polyps and the stomach were used. We proposed a stepwise process including video capture, frame extraction, pixel thresholding using both global and slice-specific values, and 3D volume rendering. The system was implemented using custom Python applications incorporating interactive threshold adjustment tools and PyVista-based 3D visualization. The workflow was tested using ultrasound videos of the gallbladder and stomach, with optional probe motion monitoring using an IMU sensor.

**Results:** The method successfully reconstructed 3D representations of gallbladder polyps. The reconstructed images provided improved spatial understanding of the anatomical relationship between lesions and surrounding structures, which were originally visible only momentarily.

**Conclusions:** This approach may serve as an initial step toward practical 3D reconstruction from routine gastrointestinal ultrasound. Further refinement and clinical validation are necessary to establish its utility in daily clinical settings.

## Introduction

Three-dimensional (3D) visualization has become increasingly important in modern diagnostic imaging, offering intuitive understanding of anatomical structures and enhancing clinical decision-making. While computed tomography (CT) and magnetic resonance imaging (MRI) routinely provide high-resolution 3D representations [1,2], ultrasound (US) remains largely limited to two-dimensional (2D) cross-sectional views in most clinical settings. In the gastrointestinal (GI) field in particular, transient lesions often appear only momentarily during scanning and can easily be missed or misinterpreted due to the lack of spatial context [3].

Two major challenges limit the use of ultrasound in dynamic 3D assessment. First, lesions may be visible for only a brief moment, making it difficult to capture or document them adequately. Second, the flexible movement of the ultrasound probe—both in position and angle relative to the skin—results in cross-sectional images whose spatial relationships are often ambiguous. It is frequently unclear which anatomical plane or slice through a 3D organ is being observed.

Various attempts have been made to extend 3D capabilities to ultrasound, including freehand volume reconstruction with position sensors [4] and motorized 3D transducers [5], but these approaches remain expensive or impractical in routine settings. More recent efforts have explored image-based 3D reconstruction without external tracking [6], though most are limited to experimental use.

In this study, we aim to establish a simple, semi-automated method for reconstructing 3D images from routine ultrasound video, with a particular focus on the gallbladder. Additionally, we explore the potential of incorporating angular velocity monitoring of the probe during scanning, using small motion sensors, to better understand and manage probe trajectory. Ultimately, we propose this method as a supportive tool to improve spatial visualization in routine GI ultrasound imaging.

## Methods

Ultrasound images were acquired using the Arietta 65 diagnostic ultrasound system (Fujifilm Healthcare Corporation, Tokyo, Japan). Ultrasound videos of author’s own gallbladder polyps and the stomach were used. For the gallbladder dataset, B-mode videos of gallbladder polyps were obtained from routine clinical examinations. For the stomach dataset, the author voluntarily underwent scanning, including upright posture and water ingestion, to simulate a full-stomach condition. Both datasets were used to test the applicability of the proposed reconstruction method under different anatomical and scanning scenarios.

### Workflow Overview

Our proposed method consists of a simple four-step process:

- **Step 1: Video Capture** Ultrasound videos were recorded when a suspected lesion became momentarily visible. Standard B-mode images were used, recorded directly from clinical equipment displays.
- **Step 2: Frame Slicing** The captured videos were converted into sequential image frames (slice images) using a custom Python script. Each frame represents a cross-sectional slice of the organ at a different probe position or angle.
- **Step 3: Thresholding** Each frame underwent threshold-based binarization. Two thresholding strategies were applied:
  - **Global threshold**: A single threshold value was applied to all slices.
  - **Slice-specific threshold**: Individual threshold values were manually adjusted for each frame using a custom threshold tuning tool (developed with Flask and OpenCV). Optionally, Canny edge detection and polygonal ROI (region of interest) trimming were used to validate boundaries and reduce noise.
- **Step 4: 3D Visualization**

Thresholded binary images were stacked and visualized as a 3D volume using PyVista, a Python-based 3D rendering library. The output mesh could optionally be exported in PLY format for further refinement.

### Probe Movement Patterns

Since precise spatial tracking of the probe was not available, we designed two simplified scanning assumptions for reconstructing the spatial structure:

- **Parallel Translation**: The probe was assumed to move linearly and perpendicularly (at 90°) to the skin surface, producing evenly spaced planar slices.
- **Fan-Shaped Movement**: The probe was assumed to pivot in a fan-like motion, simulating an arc-shaped acquisition, originally assumed to be 30–45°, though actual measurements indicated ∼15°.

These assumptions provided a geometric model for stacking and aligning the 2D slices into a 3D volume.

### Sensor Integration

To explore the integration of motion data into 3D reconstruction, we conducted a preliminary experiment using the M5StickC Plus, a compact microcontroller with an onboard IMU (inertial measurement unit) [7]. The device was temporarily mounted onto the ultrasound probe using a strap, enabling angular velocity (gyroscope) data acquisition during scanning (Fig. 1).

**Fig. 1.**
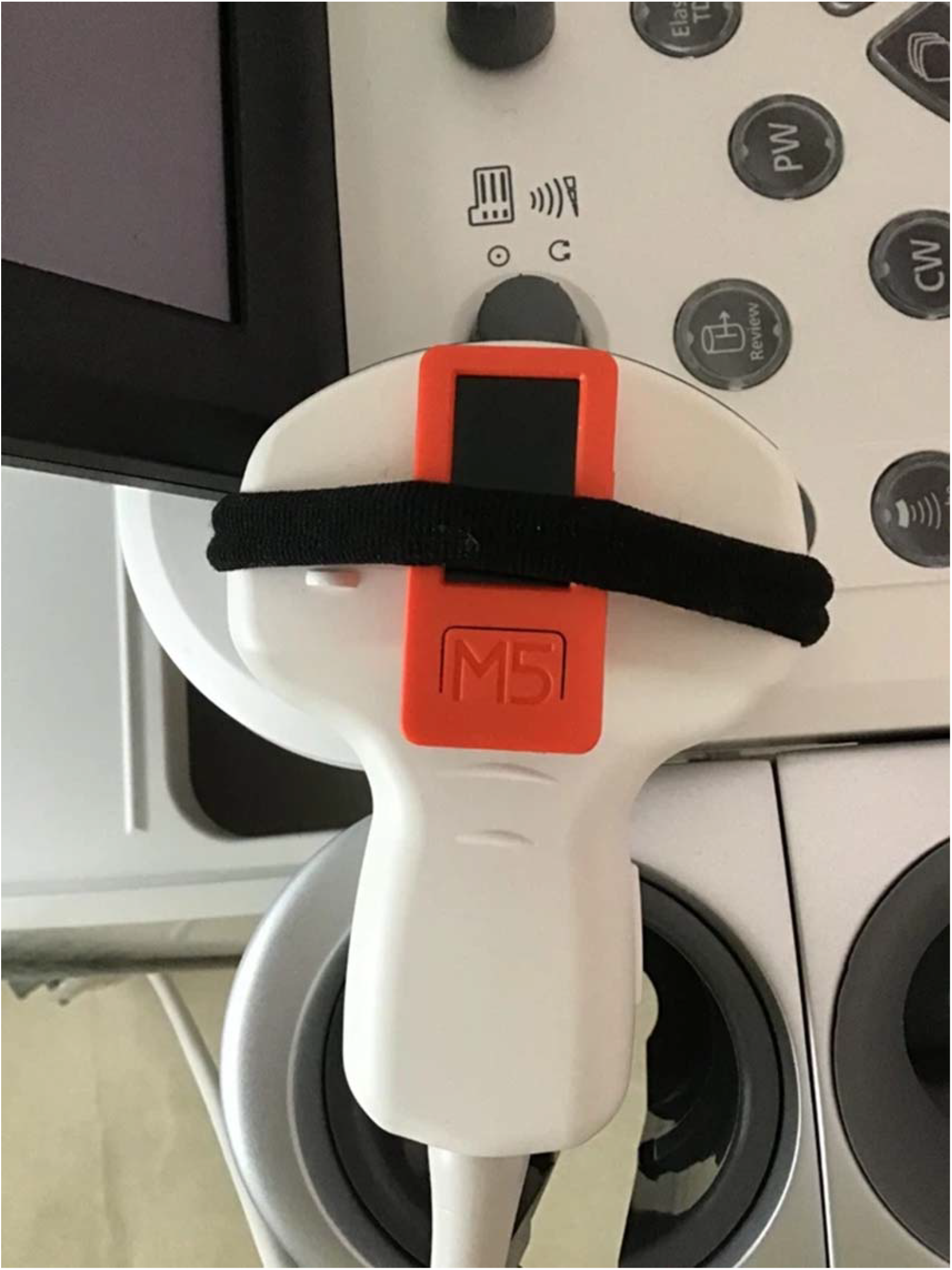
The M5StickC Plus device was temporarily mounted onto the ultrasound probe using a strap, enabling acquisition of angular velocity data via the built-in gyroscope during scanning.Although this simple setup does not provide accurate spatial localization, it demonstrates the potential of combining ultrasound images with motion data. In future implementations, high-precision probe tracking using optical markers or robotic arms may significantly improve the fidelity of 3D reconstruction.

### Tools Used (Optional)

The entire process was built using the following open-source tools and custom scripts:

- **Python & OpenCV**: For frame extraction and image thresholding.
- **Flask (Web UI)**: A lightweight server was created to allow manual threshold adjustment through a web browser interface, enabling per-frame optimization by the user.
- **PyVista**: Used for stacking 2D binary masks and rendering them into a 3D volume with real-time interaction and export functionality.
- **MeshLab**: Applied for noise reduction, hole filling, and selective mesh trimming after 3D reconstruction. The exported .ply files from PyVista were imported into MeshLab for post-processing.
- **Hardware**: An M5StickC Plus microcontroller was used to collect basic gyroscope data as a proof of concept for motion-aware reconstruction.

These tools collectively allowed the construction of a lightweight, semi-automated pipeline suitable for exploratory 3D ultrasound imaging using routine clinical video data.

## Results

### Overview of the Workflow

Our proposed 3D reconstruction pipeline consists of four steps: video capture during routine abdominal ultrasound, frame extraction, binarization via thresholding (global or per-slice), and 3D rendering using PyVista. MeshLab was optionally used to refine the surface quality of the reconstructed mesh (Table 1).

**Table I.** Summary of Workflow Steps.

| Step | Description | Tools Used |
| --- | --- | --- |
| 1 | Capture ultrasound video | Ultrasonic machine, MP4 output |
| 2 | Extract sequential frames from video | mp4_to_jpg_with_time.py |
| 3 | Apply thresholding to each frame | pixel_point.py,<br>generate_threshold_masks.py |
| 4 | Render 3D volume from processed slices | main_pyvista_3d.py |
| 5 | (Optional) Mesh editing and refinement | 3DSliceViewer_with_ply_export.py,<br>MeshLab |

### Gallbladder Polyp Reconstruction

A B-mode ultrasound video of the gallbladder was processed using the proposed workflow. From the original MP4 file, 150 JPEG frames were extracted. Threshold values between 3 and 20 were determined interactively using a custom-built GUI (Fig. 2–5). The resulting point cloud was converted into a 3D mesh and visualized with volume rendering. Further refinement with MeshLab enabled removal of artifacts and improved surface smoothness (Fig. 6).

**Fig. 2.**
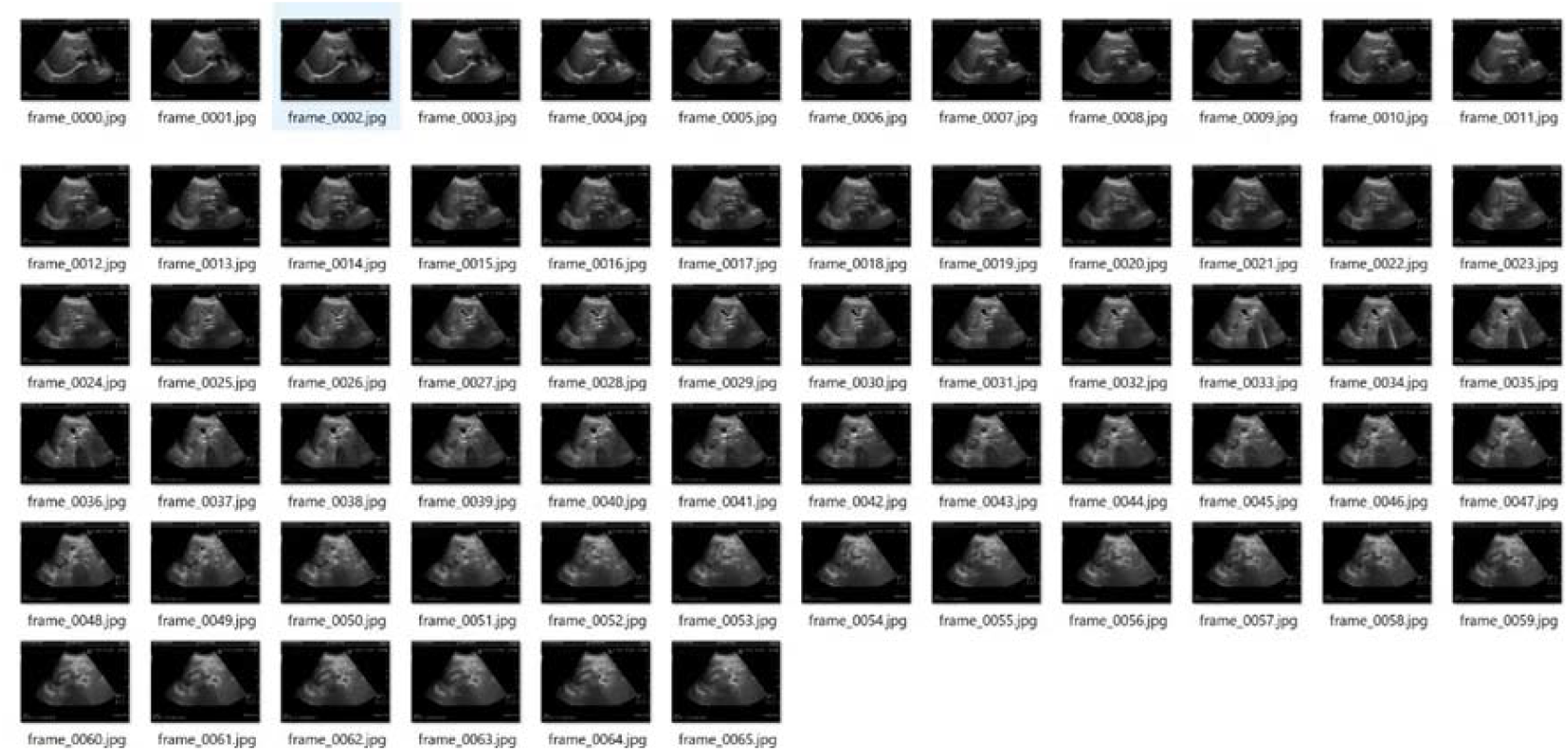
Example of frame extraction using the script “mp4_to_jpg_with_time.py.”

**Fig. 3.**
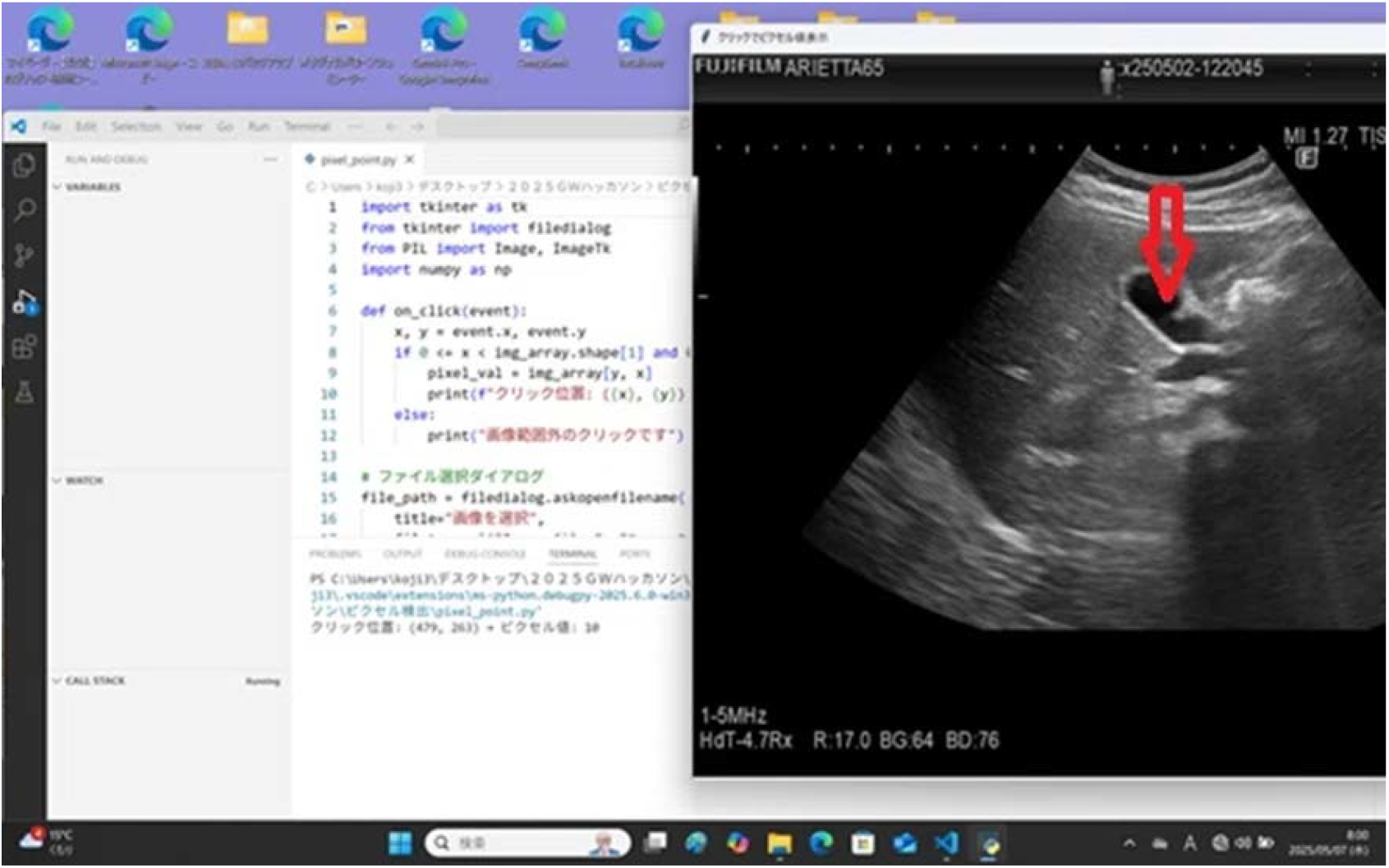
Screenshot of the threshold selection interface implemented in “pixel_point.py.”

**Fig. 4.**
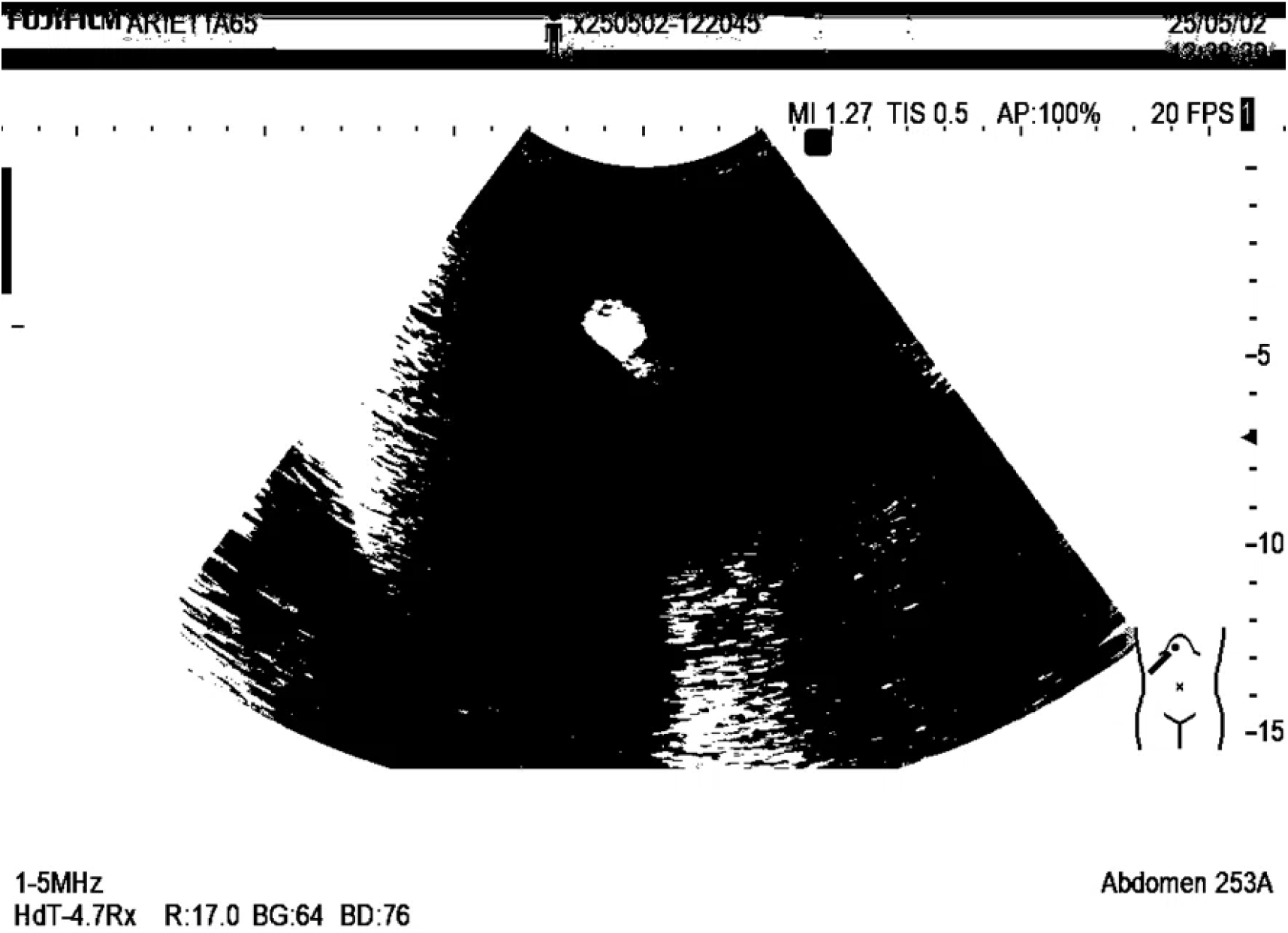
Result of applying a global threshold to all extracted frames.

**Fig. 5.**
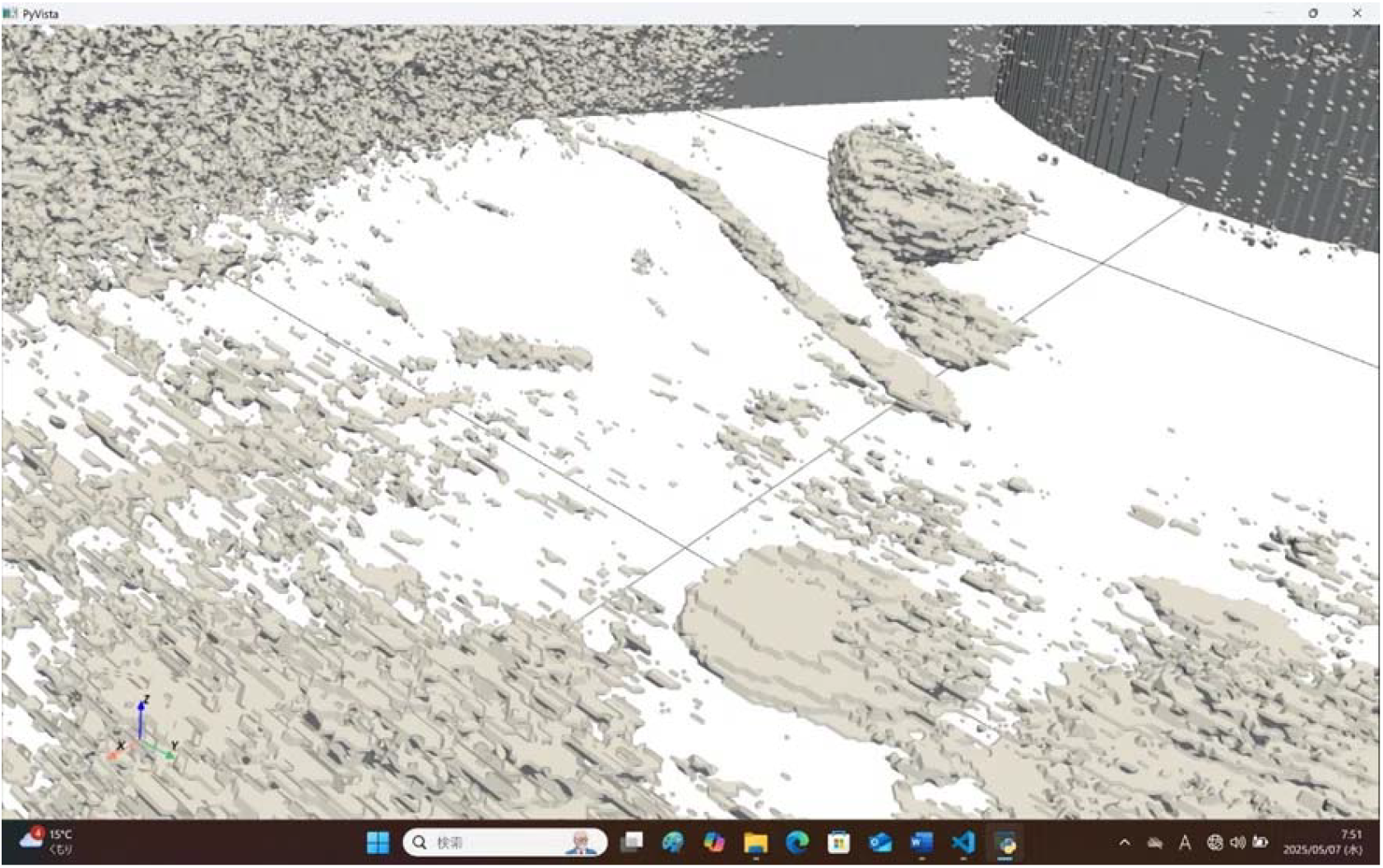
Three-dimensional reconstruction of a gallbladder polyp using PyVista.

**Fig. 6.**
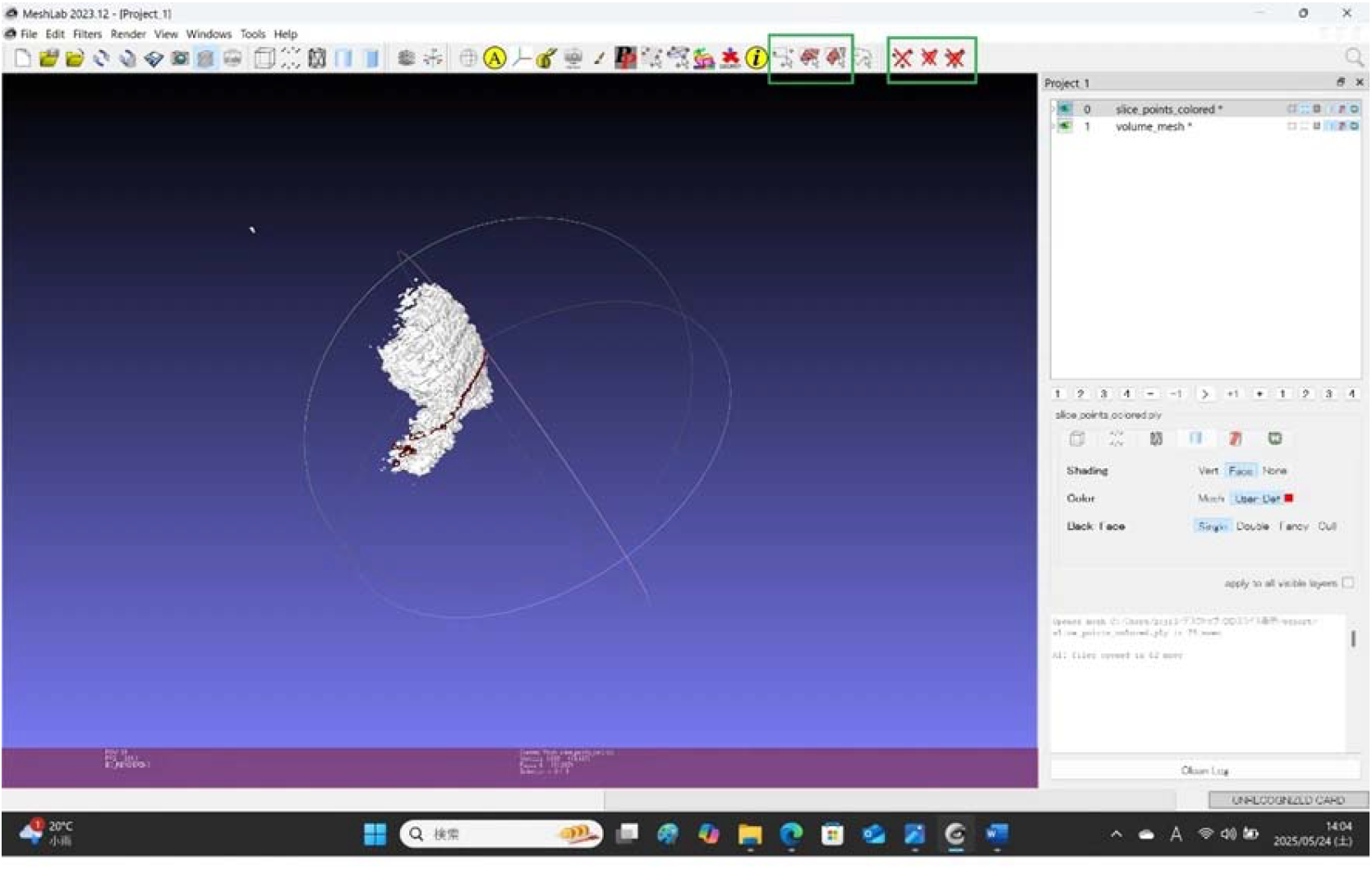
Post-processed 3D rendering of the gallbladder polyp, refined using MeshLab.

### Gastric Imaging (Preliminary Application)

We also applied the workflow to an ultrasound video of the stomach acquired in a standing position after water ingestion. The fan-shaped movement of the probe was estimated to span approximately 30°, although no quantitative tracking was performed (Fig. 7).

**Fig. 7.**
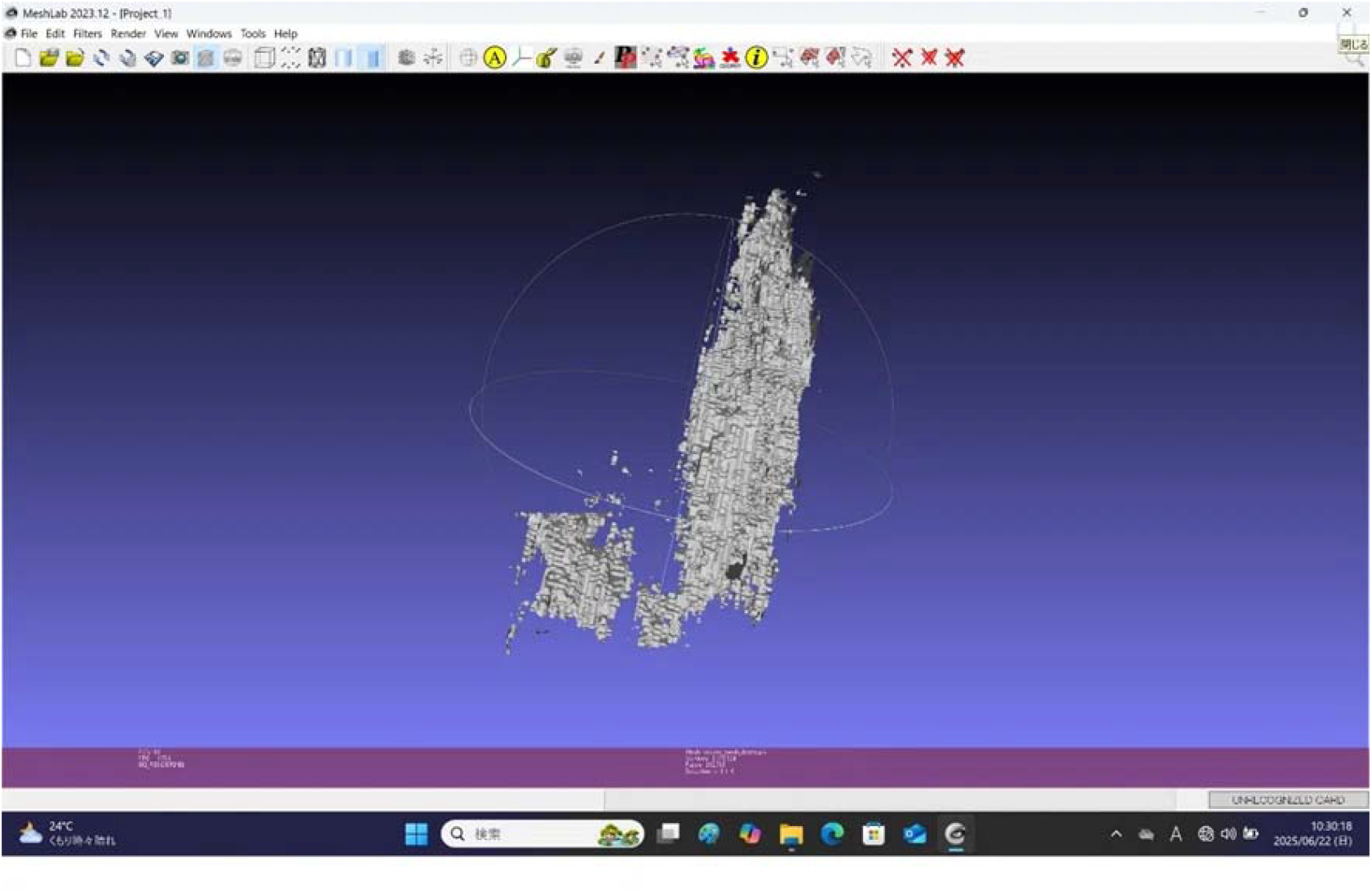
Three-dimensional image of the stomach constructed from ultrasound frames and refined with MeshLab.

### Effect of Different Probe Motion Assumptions

To evaluate the influence of probe motion assumptions on reconstruction fidelity, we generated three 3D models of the same gallbladder polyp using different motion parameters:

1. Linear translation over 60 mm in 6 seconds.
2. A theoretical 45° fan-shaped sweep.
3. A measured 15° sweep using an IMU (M5StickC Plus).

All reconstructions were performed using identical frame sequences and thresholds. The linear and 45° fan-shaped models resulted in comparable gallbladder shapes, whereas the 15° model yielded a slightly compressed morphology, potentially reflecting the true anatomical structure. (data not shown)

## Discussion

This study presents a novel, semi-automated workflow for reconstructing three-dimensional (3D) representations from routine ultrasound videos, focusing primarily on gallbladder polyps and, to a lesser extent, gastric anatomy. While 3D imaging has become standard in CT and MRI, its implementation in ultrasonography remains rare due to challenges such as non-uniform probe motion, limited spatial cues, and the inherently two-dimensional nature of ultrasound imaging.

### Clinical Relevance and Novelty

To date, only a limited number of studies have reported practical 3D reconstructions from gastrointestinal ultrasound data [8,9]. Our method addresses a key diagnostic gap: the inability to spatially contextualize lesions that appear only momentarily during scanning. By enabling post hoc 3D visualization from standard B-mode videos, the proposed system enhances the spatial understanding of lesion location, depth, and anatomical relationships—particularly valuable in hepatobiliary imaging.

### Technical Contributions

The pipeline was developed using open-source Python tools. It includes sequential frame extraction, manual or slice-specific thresholding via a Flask-based GUI, and volumetric rendering using PyVista. Optional boundary refinement with Canny edge detection and 3D mesh smoothing with MeshLab further enhanced image quality. This modular approach provides flexibility and scalability for different clinical applications.

Furthermore, the open-source platform **3D Slicer** [10] represents a promising foundation for future integration, offering real-time 3D visualization and extensible plugin architecture suitable for clinical deployment.

### Limitations and Motion Tracking

Despite promising results, several limitations remain. First, manual threshold adjustment for each slice is time-consuming and introduces user-dependent variability. Future versions should explore adaptive or AI-based thresholding methods to improve consistency and speed.

Second, gastric imaging poses inherent challenges: even after water ingestion, insufficient gastric distension and low contrast often result in partial or collapsed wall reconstructions.

Third, our reconstruction currently assumes simplified probe trajectories—linear or fan-shaped—which may not accurately reflect the complex motion used during real clinical scanning.

To address this, we implemented a preliminary motion analysis using a compact inertial measurement unit (IMU), the **M5StickC Plus**. While the operator estimated a 45° sweep during fan-shaped scanning, IMU data revealed an actual angular movement of approximately 15°. This discrepancy underscores the limitations of visual estimation and emphasizes the need for objective probe motion tracking.

Interestingly, 3D reconstructions generated under three different assumptions—linear translation, theoretical 45° arc, and IMU-measured 15° arc—revealed subtle morphological differences, with the IMU-based model producing a slightly compressed gallbladder shape that may more closely represent the true anatomy (data not shown). The feasibility of integrating such low-cost IMUs into biomedical workflows is supported by prior research on human motion tracking and rehabilitation monitoring [11,12].

### Comparison and Future Directions

Future work should position this technique alongside established volumetric imaging methods such as **CT colonography** and MRI-based reconstruction, which remain the gold standards for 3D gastrointestinal visualization. Notably, CT colonography has demonstrated high diagnostic accuracy in detecting colorectal neoplasia in asymptomatic adults [13]. However, ultrasound provides distinct advantages—real-time imaging, portability, and absence of ionizing radiation—that make it ideal for bedside applications and repeated follow-up examinations.

To improve geometric accuracy and reproducibility, future systems should integrate robust motion-tracking frameworks, including synchronized IMU-GPS systems, optical tracking, or robotic probe holders [14]. These technologies could allow real-time fusion of probe motion data with image slices, enabling automated correction of spatial misalignment during 3D stacking and rendering.

In addition, automatic lesion segmentation and adaptive thresholding algorithms will be critical to streamline workflow and reduce operator dependency. Software ecosystems such as 3D Slicer, which support real-time visualization, plugin development, and DICOM interoperability, may help bridge the gap between research prototypes and clinical deployment.

### Educational and Broader Applications

Beyond diagnostic use, the proposed system offers significant educational potential. Interactive 3D reconstructions generated from conventional ultrasound allow medical students and trainees to intuitively grasp anatomical relationships and spatial orientation. This approach can thus serve as a valuable training resource, supporting both clinical education and research in ultrasonographic imaging.

## Conclusion

This preliminary study demonstrates the feasibility of three-dimensional reconstruction from routine ultrasound videos using a semi-automated workflow, primarily applied to the gallbladder. By combining frame slicing, tailored image thresholding, and volumetric rendering with open-source tools, we were able to create intuitive 3D representations of transient lesions that are otherwise difficult to interpret in conventional two-dimensional ultrasound.

The proposed approach offers a new direction for enhancing the spatial understanding of gastrointestinal anatomy, especially in cases where lesions appear only momentarily. While current limitations—such as manual threshold adjustment and untracked probe motion—remain, the methodology serves as a promising foundation for future development.

With further refinement and clinical validation, including integration of sensor-based probe tracking and AI-driven segmentation, this technique has the potential to expand the diagnostic capabilities of abdominal ultrasound and pave the way toward practical 3D ultrasonographic tools in daily medical practice.

## Disclosures

The author declares no conflicts of interest related to this study.

All procedures performed in this study involving human participants were conducted in accordance with the ethical standards of the institutional research committee and with the 1964 Helsinki Declaration and its later amendments. Informed consent was obtained from all individual participants included in the study.

This article does not contain any studies with animals performed by the author.

## Code and Data Availability

The data and code supporting the findings of this study are available from the corresponding author upon reasonable request. The video and image data used in this study were obtained from the author’s own routine ultrasound examinations and are not publicly available due to privacy considerations. The custom Python scripts used for semi-automated 3D reconstruction are available upon request for academic, non-commercial purposes.

